# A community-based validation of the International Alliance for the Control of Scabies Consensus Criteria by expert and non-expert examiners in Liberia

**DOI:** 10.1101/2020.06.08.20125682

**Authors:** SL Walker, S Collinson, J Timothy, S Zayay, K Kollie, E Lebas, K Halliday, R Pullan, M Fallah, M Marks

## Abstract

**Background:** The International Alliance for the Control of Scabies (IACS) recently published expert consensus criteria for scabies diagnosis. Formal validation of these criteria is needed to guide implementation. We conducted a study to provide detailed description of the morphology and distribution of scabies lesions as assessed by dermatologists and validate the IACS criteria for diagnosis by both expert and non-expert examiners.

**Methods:** Participants from a community in Monrovia, Liberia, were independently assessed by two dermatologists and six mid-level healthcare workers. Lesion morphology and distribution were documented based on the dermatologist examination. Diagnoses were classified by IACS criteria and the sensitivity and specificity of MLHW assessments calculated.

**Results:** Papules were the most common lesions (97.8%). Burrows were found in just under half (46.7%) and dermatoscopy was positive in a minority (13.3%). Scabies lesions were found in all body regions but more than 90% of patients could have been diagnosed by an examination of only the limbs. Severity of itch was associated with lesion number (p=0.003). The sensitivity of MLHWs to detect typical scabies ranged between 69-83% and specificity 70-96%. The sensitivity of MLHWs was higher in more extensive disease (78-94%).

**Conclusions:** The IACS criteria proved a valid tool for scabies diagnosis. For the purposes of implementation papules and burrows represent truly ‘typical’ scabies lesions. MLHWs are able to diagnose scabies with a high degree of accuracy, demonstrating they could form a key component in population-level control strategies.

**Plain English Summary:** Scabies is a very common skin condition in both high- and low-income settings with hundreds of millions of people affected each year. Recently standardised criteria have been proposed to help improve the quality of scabies diagnosis, in particular in low income settings where the access to a skin specialist is very limited.

In this study, conducted in Liberia, expert examiners conducted a thorough examination and recorded what different types of skin problems they found in participants with and with and without scabies. We then compared the accuracy of a diagnosis of scabies made by dermatologists to that made by non-specialist healthcare workers who had received a short training course over three days.

We found that small bumps (papules) were the most common type of scabies lesion and were found in almost every single patient with scabies. A second type of skin lesion called a burrow was the next most common and was found in just under half of the participants. Other types of scabies lesions which have been described were rare in this study. We found that after the short training course the non-specialists were able to detect about the majority of the cases of scabies correctly.

Our study has helped provide detailed data on exactly what types of skin changes are typical of scabies and demonstrated how short training programmes can help improve the skill of non-specialist examiners in diagnosing scabies.

## Introduction

Scabies is a severe pruritic skin disease caused by the mite *Sarcoptes scabei* var *hominis*, which is a significant public health problem in many low-income settings. Globally, there are believed to be more than 400 million cases of scabies each year[1] and it is one of the commonest dermatoses that a health care provider will encounter in low-income settings [2].

The mainstay of diagnosing scabies is a thorough history and detailed clinical examination. Clinical examination may be complemented by other techniques including dermatoscopy, non-invasive higher power imaging devices or light microscopy [3] of skin specimens which allow definitive parasitological diagnosis. However, they have low sensitivity, are time consuming and are impractical in many low-income settings due to financial and personnel constraints[3]. In most low-income settings there is an absence of trained individuals with expertise in skin disease and health systems are dependent on mid-level health workers (MLHWs) such as clinical officers to diagnose and manage patients with skin disease.

The adoption of scabies as a Neglected Tropical Disease (NTD) by the World Health Organization (WHO) has led to the development of scabies control programmes which would benefit from robust methods of diagnosis. Strategies have previously been developed and validated to aid MLHWs in the diagnosis of scabies, impetigo and other common dermatoses. These approaches have been shown to have acceptable sensitivity and specificity when compared to examination by a reference standard expert examiner [4–7]. Whilst promising, one challenge has been the lack of validated diagnostic criteria for scabies. The International Alliance for the Control of Scabies (IACS) [8] developed and recently published a detailed description of diagnostic criteria [3,9] for three levels of diagnostic certainty: ‘A - Confirmed Scabies’ which requires visualisation of the mite, ova or scybala; ‘B-Clinical Scabies’ and ‘C - Suspected Scabies’.

In addition to the presence of scabies burrows or typical lesions affecting the male genitalia the 2020 IACS criteria for Clinical Scabies include ‘typical lesions’ in a ‘typical distribution’ in an individual with itch and a history of contact with someone with scabies or someone with unexplained itch. The ‘typical lesions’, other than burrows, are defined as papules, nodules, vesicles and pustules. A typical distribution in adults is defined as lesions affecting one or more of the following sites: the distal forearm and hands, the axilla, the umbilical region, the groin and legs whilst in infants (children under the age of two) all body sites may be affected [3]. A complete skin examination is recommended but more limited examinations may still have a high diagnostic yield and are more practical in non-clinical settings [10].

We conducted a prospective study to validate the performance of the IACS criteria by both expert and trained non-expert examiners including a detailed description of the lesion morphology and distribution in individuals with scabies.

## Materials and methods

This was a prospective diagnostic accuracy study conducted in urban Monrovia, Liberia in February 2020.

### Training of non-expert examiners

Training was delivered by two specialists in dermatology with experience of managing skin disease in low income settings (EL and SW). Five MLHWs participated in the two-day training workshop.

The first day of training consisted of classroom-based tutorials on the morphology of skin lesions and the clinical features and treatment of scabies, impetigo, infected scabies and dermatophyte infections. The second day consisted of supervised clinical training in People’s United Community (PUC), Sinkor district, where the MLHWs performed clinical skin examination and made diagnoses.

### Validation study

We conducted a validation exercise with the two specialists, the five recently trained MLHWs and an additional MLHW who had received training 18 months earlier as part of a separate study on screening for Buruli ulcer, leprosy, yaws and lymphatic filariasis but had not received the most recent training. Residents from the Raymond Field/Barrolle Practice Ground community, Sinkor district, were invited to attend for assessment and treatment of skin problems. Each participant was examined independently by the MLHWs and both specialists in a private setting.

The MLHWs recorded whether a participant had itch and/or a history of contact with an individual with itch. They performed a skin examination (excluding the genitals) and recorded the presence of a skin problem and whether skin lesions were typical in morphology and distribution for scabies. They recorded the number of scabies lesions (1-10, 11-49 or ≥50) to assess the extent of disease [11,12]. In individuals diagnosed with scabies, the MLHWs recorded the presence of any secondary bacterial infection and classified the number of infected lesions: 1-5, 6-10, 11-49, ≥50.

The two specialist examiners also elicited itch and contact history and each participant was asked about severity of itch using the Severity of Pruritus Scale[13]. A clinical examination with the aid of a dermatoscope (Heine Delta 20 Plus, Herrsching, Germany) was performed. Dermatoscopic findings for scabies were recorded as positive or negative. Positive dermatoscopy was categorised as visualisation of the mite and the presence of the ‘wake’ sign was also recorded [3]. For the purpose of IACS classification only the former was considered diagnostic of confirmed scabies. Individuals diagnosed with scabies had the morphology of lesions (papules, vesicles, nodules and burrows) and number of each at 19 pre-defined body sites recorded by one examiner (SW). The consensus diagnosis of the two expert examiners was used as a reference standard with which to evaluate the performance of the non-expert examiners. The sensitivity and specificity of each non-expert examiner compared to the reference standard was calculated and their diagnoses were classified by IACS category B1, B3, C1 or C2. MLHWs did not document specific lesion sites or examine the genitals and thus an assessment of their performance in using category B2 (male genital lesions) was not possible.

It was calculated that at least 40 individuals with and without scabies were needed to detect a sensitivity of the non-expert examiners of 90% +/-10% compared to the reference standard.

Data were collected anonymously directly on Android devices (Samsung Galaxy Tab A) using the Open Data Kit (ODK, Seattle, USA, 2010) application and uploaded remotely to the dedicated secure server at the London School of Hygiene and Tropical Medicine. Data were analysed using R 3.3.0[14]. Individuals who were diagnosed with scabies by the expert examiners were offered treatment with oral ivermectin or benzyl-benzoate lotion.

Ethical approval was obtained from the Ethics Committees of the London School of Hygiene and Tropical Medicine (Reference 17796) and the University of Liberia – Pacific Institute for Research and Evaluation Institutional Review Board (Reference 20-01-195). Written informed consent was obtained from participants aged 18 years and older and from the parents or guardians of children. Verbal assent was obtained from children who were able to provide it.

## Results

One hundred and forty-seven individuals were examined by the expert examiners and of these 135 were examined by all the non-expert examiners and were included in the validation analyses. The 12 participants who were not examined by all the non-expert examiners were excluded from calculations of the sensitivity and specificity of MLHWs for the diagnosis of scabies.

### Diagnoses of overall cohort

One hundred and forty-seven participants underwent examination by the expert examiners. The median age was 17 years (IQR 6-31) and 97 participants (70%) were female. 128 individuals (87.1%) had a cutaneous diagnosis and 139 individuals (94.6%) reported a history of itch. Scabies was the commonest diagnosis; 44 individuals (29.9%) were diagnosed with scabies by both expert examiners. There were a further two cases where the expert examiners disagreed about a diagnosis of scabies (1.4%). Only four individuals (2.7%) had infected lesions. The median age of participants with scabies was 11 (IQR 3 -23) and the majority were females (n = 28, 64.4%). The other two most common diagnoses were dermatophyte infections (n = 23, 15.6%) and atopic dermatitis (n =17, 11.6%) (Table 1).

**Table 1:** Diagnoses made on examination by a dermatologist.

| Diagnosis | n (%) |
| --- | --- |
| Scabies | 45 (30.6%) |
| Tinea corporis | 15 (10.3%) |
| Atopic dermatitis/Eczema | 17 (11.5%) |
| Lichen Simplex | 9 (6.2%) |
| Tinea capitis | 7 (4.8%) |
| Folliculitis | 6 (4.1%) |
| Follicular Eruption | 6 (4.1%) |
| Impetigo | 4 (2.8%) |
| Pityriasis versicolor | 4 (2.8%) |
| Acne | 3 (2.1%) |
| Other* | 21 (14.3%) |
| No skin problem | 19 (12.9%) |

\*Details of conditions grouped under other are given in Supplementary Table 3

### Clinical Features of the IACS criteria assessed by an expert examiner

Forty-five of the forty-six cases of scabies diagnosed by either expert examiner underwent a comprehensive examination including full body examination, lesion counting and dermatoscopy. The IACS classification for these 45 individuals is shown in Table 2. Papules were the most common lesion type and were present in 45 individuals (97.8%) with scabies followed by burrows (n = 21, 46.7%) (**Table 2**). The median number of scabies lesions was 117 (IQR-47-164) of which the majority were papules (**Table 2**). All individuals with scabies reported a personal history of itch and 43 (93.5%) reported a history of contact with an individual with itch. Dermatoscopy was consistent with scabies in 19 individuals, all of whom had burrows on examination and mites were visualised in 6 of these cases (**Table 2**). The number of scabies lesions was strongly associated the degree of reported itch (p = 0.003) graded using the Severity of Pruritus Scale. Individuals with mild itch had a median of 55 lesions (IQR 46-77), those with moderate itch a median of 98 lesions (IQR 41-148) and a median of 159 lesions (IQR 133-256) was found amongst individuals who reported severe itch with sleep disturbance (**Figure 1**).

**Table 2:** Detailed clinical features of scabies on comprehensive examination by a dermatologist.

| Clinical Features |  |  |
| --- | --- | --- |
| IACS Classification | A3: Dermatoscopy Confirmed | 6 (13.3%) |
|  | B1: Presence of Burrows | 15 (33.3%) |
|  | B2: Typical Genital Lesions | 3 (6.7%) |
|  | B3: Typical Lesions in a Typical Distribution with 2 History Features | 19 (42.2%) |
|  | C1: Typical Lesions in a Typical Distribution with 1 History Features | 2 (4.4%) |
| Lesion Type Present | Papule | 44 (97.8%) |
|  | Burrow | 21 (46.7%) |
|  | Nodule | 12 (26.7%) |
|  | Vesicle | 1 (2.2%) |
| Lesion Type Count (Median, IQR) | Papule | 115 (45-161) |
|  | Burrow | 0 (0-3) |
|  | Nodule | 0 (0-1) |
|  | Vesicle | 0 (0-0) |
| Itch | Personal History | 45 (100%) |
|  | Household Contact | 39 (86.7%) |
|  | Non-Household Contact | 17 (37.8%) |
| Dermatoscopy | Delta Wing & Wake Sign | 4 (8.7%) |
|  | Delta Wing Sign Only | 2 (4.3%) |
|  | Wake Sign Only | 13 (28.2%) |

\*All participants with dermatoscopic confirmation of scabies also had burrows

**Figure 1:**
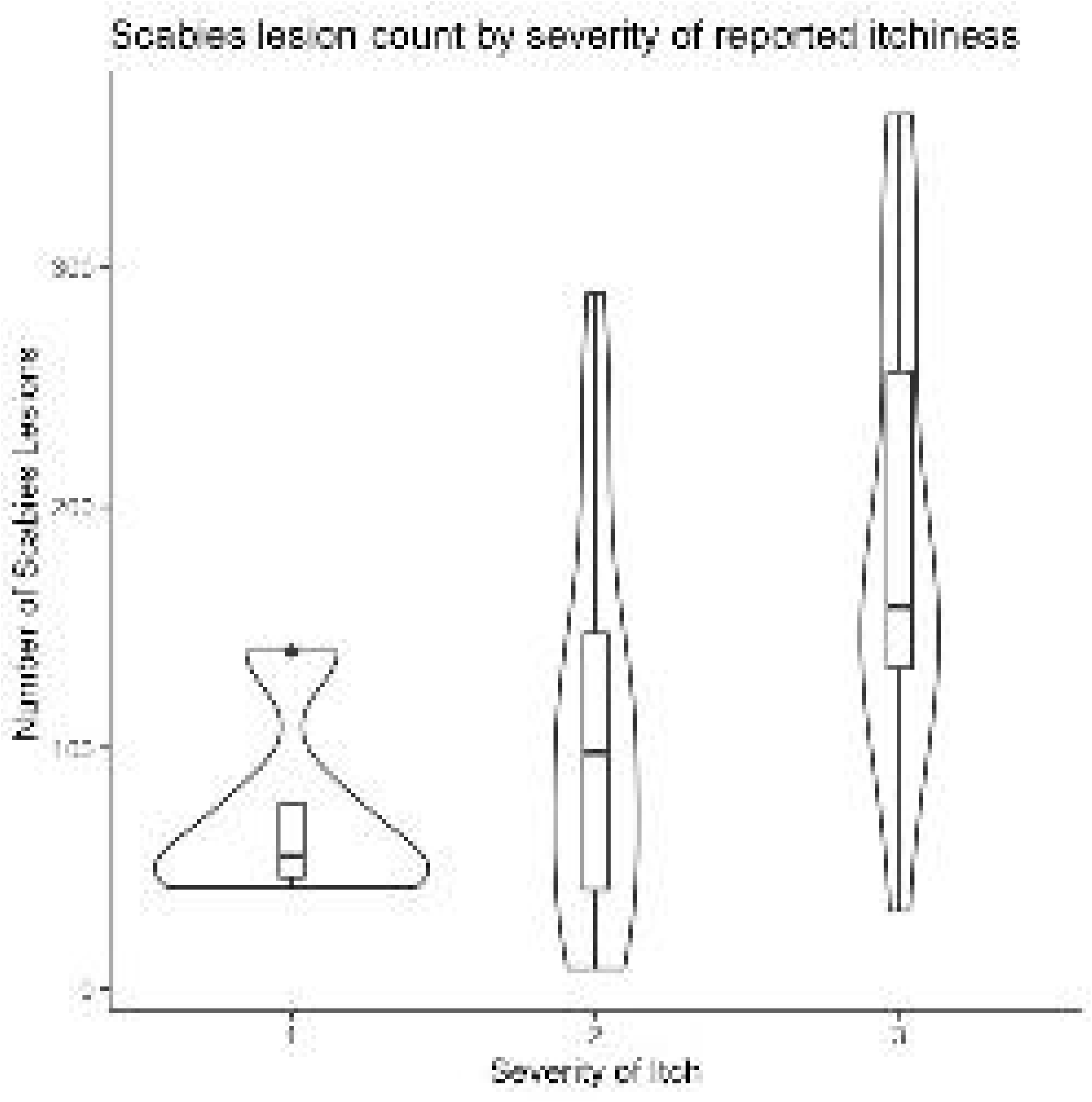
Number of scabies lesions in relation to grade of reported severity of itch using the Severity of Pruritus Scale.

The buttocks and groin (76%), wrist (73%), torso (71%), forearm (67%), and inter-digital web-spaces (64%) were the most common location for papules (**Table 3**). Compared to a full body examination for all lesion types, papules could have been detected through a limited examination involving the face, and upper limb including the axilla and fingers in 91.1% of cases of scabies.

**Table 3:** Proportion of individuals with scabies with lesions in different body regions (n=45)

| Body Region | PAPULES | BURROWS | NODULES | VESICLES |
| --- | --- | --- | --- | --- |
| Face and neck | 13.3% (6) | 0% | 0% | 0% |
| Torso (excluding peri-areola and peri-umbilicus) | 71.1% (32) | 2.2% (1) | 4.4% (2) | 2.2% (1) |
| Peri-areola | 24.4% (11) | 0% | 0% | 0% |
| Peri-umbilicus | 40.0% (18) | 0% | 4.4% (2) | 0% |
| Axillary vault | 42.2% (19) | 0% | 0% | 0% |
| Arm | 40.0% (18) | 0% | 0% | 0% |
| Elbow | 35.6% (16) | 0% | 4.4% (2) | 0% |
| Forearm | 66.7% (30) | 0% | 4.4% (2) | 0% |
| Wrist | 73.3% (33) | 24.4% (11) | 0% | 0% |
| Palm of hand | 26.7% (12) | 17.8% (8) | 0% | 0% |
| Dorsum of hand | 62.2% (28) | 2.2% (1) | 0% | 0% |
| Inter-digital web spaces | 64.4% (29) | 8.9% (4) | 0% | 0% |
| Fingers | 42.2% (19) | 0% | 0% | 0% |
| Buttocks and groin | 75.6% (34) | 0% | 15.6% (7) | 0% |
| Male genitalia (penis and scrotum)* | 62.5% (10) | 18.8% (3) | 0% | 0% |
| Thigh (including knee) | 60% (27) | 0% | 15.6% (7) | 0% |
| Leg (including ankle) | 57.8% (26) | 0% | 4.4% (2) | 0% |
| Dorsum of foot | 33.3% (15) | 0% | 0% | 0% |
| Sole of foot | 8.9% (4) | 6.7% (3) | 0% | 0% |

\*Amongst male participants only

### Performance of Non-Expert Examiners

The median age of the 135 participants examined by all the MLHWs was 18 years (IQR 7-32) and 89 (65.9%) were female. The expert examiners reached a consensus diagnosis of scabies in 42 individuals (31.1%) which included all four cases of infected scabies.

When we considered any of the categories B1, B3, C1 or C2 as diagnostic of scabies the sensitivity of the non-expert examiners ranged between 73% - 93% and the specificity ranged between 56% and 96%. When we excluded IACS Category C2 (presence of either atypical lesions or an atypical distribution) the sensitivity of the non-expert examiners ranged between 69-83% and the specificity between 70-96% (**Table 4**). The sensitivity of non-expert examiners was lower in scabies affected individuals with fewer lesions (range 30-60%) and higher in more extensive disease (range 78-94%). The six MLHWs made 160 false positive diagnoses. 104 of 160 (65%) of these false positive diagnoses were accounted for by atopic dermatitis/eczema, folliculitis, tinea corporis and capitis, lichen simplex, lichen planus and pityriasis versicolor. These seven pruritic conditions (of the 25 non-scabies diagnoses made) accounted for 65.2% of people with non-scabies diagnoses.

**Table 4:** Performance of Non-expert examiners.

|  |  | IACS Category |  |  |  |  | B1-B3 and C1-C2 |  |  |  | B1-B3 and C1 |  |  |  |
| --- | --- | --- | --- | --- | --- | --- | --- | --- | --- | --- | --- | --- | --- | --- |
|  |  | B1 | B2 | B3 | C1 | C2 | Sensitivity %<br>(95% CI) | Specificity%<br>(95% CI) | PPV%<br>(95% CI) | NPV%<br>(95% CI) | Sensitivity<br>% (95% CI) | Specificity<br>% (95% CI) | PPV%<br>(95% CI) | NPV% (95%<br>CI) |
| Non-Expert Examiner | 1 | 3 | 0 | 25 | 5 | 2 | 73.8<br>(57.7-85.6) | 95.7<br>(88.7 – 98.6 ) | 88.6<br>(72.3-96.3) | 89<br>(80.8-94.1) | 69.1<br>(52.8-81.9) | 95.7<br>(88.7 – 98.6 ) | 87.9<br>(70.9-96.0) | 87.3<br>(78.8-92.8) |
|  | 2 | 20 | 0 | 20 | 8 | 13 | 83.4<br>(68.0-92.5) | 72.1<br>(61.6-80.6) | 57.4<br>(44.1-69.7) | 90.5<br>(80.9-95.8) | 76.2<br>(60.2-87.4) | 82.8<br>(73.3-89.6) | 66.7<br>(51.5-79.2) | 88.6<br>(79.4-94.1) |
|  | 3 | 0 | 0 | 49 | 6 | 23 | 92.9<br>(79.4-98.1) | 57<br>(46.3-67.1) | 53.5<br>(41.4-65.0) | 94.7<br>(84.2-98.6) | 78.6<br>(62.8-89.2) | 76.4<br>(66.2-84.3) | 60<br>(45.9-72.7) | 88.8<br>(79.2-94.4) |
|  | 4 | 3 | 0 | 40 | 13 | 17 | 92.9<br>(79.4-98.1) | 63.5<br>(52.8-73.0) | 53.5<br>(41.4-65.0) | 95.2<br>(85.6-98.7) | 78.6<br>(62.8-89.2) | 75.3<br>(65.0-83.4) | 58.9<br>(45.0-71.6) | 88.6<br>(79.0-94.3) |
|  | 5 | 0 | 0 | 33 | 10 | 5 | 78.6<br>(62.8-89.2) | 83.9<br>(74.5-90.4) | 68.8<br>(53.6-80.9) | 89.7<br>(90.8-94.9) | 76.2<br>(60.2-87.4) | 88.2<br>(79.4-93.7) | 74.5<br>(58.5-86.0) | 89.1<br>(80.5-94.4) |
|  | 6 | 1 | 0 | 52 | 10 | 16 | 90.5<br>(76.5-96.9) | 56<br>(45.3-66.1) | 48.1<br>(36.8-59.6) | 92.9<br>(81.9-97.7) | 83.3<br>(68.0-92.5) | 69.9<br>(59.4-78.7) | 55.6<br>(42.6-67.9) | 90.3<br>(80.4-95.7) |

The non-expert examiner trained 18 months previously and who attended only for the validation study was equivalent to the non-expert examiners who had undergone training and assessment in the week of the study (**Table 4**).

## Discussion

This is the first study to undertake validation of the 2020 IACS Consensus Criteria for the Diagnosis of Scabies, which incorporate a comprehensive explanation of how to apply the diagnostic criteria. We found that agreement between the two expert examiners on the presence of scabies was high (96% of cases), providing confidence in our reference standard diagnosis.

The symptoms and signs used to define five (A3, B1, B2, B3 and C1) of the six categories tested showed diagnostic validity in this setting with a complete skin examination performed by dermatologists and supplemented with dermatoscopy. The diagnostic components with highest sensitivity for scabies were the history components, a personal history and a contact history, present in 100% and 93.5% of cases respectively and papular lesions which were present in 97.8%. Our results highlight that papular lesions are the predominant lesion type in scabies. Given our findings it may be reasonable in this setting to define ‘typical lesions’ as papular lesions or burrows with neither nodules nor vesicles contributing significantly. The expert examiners performed a comprehensive skin examination which allowed us to calculate that more than 90% of cases could possibly have been detected by a more limited examination of the face, upper limb including the axilla and fingers. The sensitivity of this limited examination supports its utility as a practical alternative for use in large scale field settings such as community-wide surveys[10].

MLHWs were able to diagnose scabies with a high degree of accuracy after attending a two-day training programme compared with the reference standard diagnosis made by two skin specialists. When including IACS 2020 diagnostic criteria for clinical and suspected scabies categories B1, B3 and C1, C2, the mean sensitivity of MLHW diagnosis was high at 85.4% (range 73-93%), with a specificity of 71.4% (range 56-96%). In LMIC settings there are few trained skin specialists, but our results indicate that in the absence of such experts and specialist diagnostic equipment, trained MLHWs are able to diagnose scabies [3]. An MLHW who had received training on the diagnosis of scabies 18 months previously performed to a comparable standard to more recently trained MLHWs. This suggests that diagnostic skills can be retained which would greatly strengthen the benefits of appropriately evaluated, structured training programmes for MLHWs. However, this requires further assessment.

The sensitivity achieved by the MLHWs in this study is higher than that reported in a comparable study examining the diagnostic accuracy of non-expert examiners in the Solomon Islands, which documented a sensitivity of 55.3% (range 41.5-64.9%)[11]. The authors also reported that the sensitivity of non-experts in this study was higher for more extensive scabies and lower in those with fewer skin lesions. Only including the IACS diagnostic criteria that consider typical lesions or distributions (i.e. excluding category C2: presence of either atypical lesions or an atypical distribution), the mean sensitivity of MLHW diagnosis was moderately lower at 77% (range 69-93%) but specificity increased to a mean of 81.4% (range 70-96%). This suggests, perhaps unsurprisingly, that MLHW diagnosis of scabies is more accurate when patients present with typical features. Future training of MLHWs would likely be improved by including training on recognition of common dermatoses which confound the diagnosis of scabies as these contribute significantly to false positive diagnoses.

We have shown that the utility of the C2 categorisation of suspected scabies does not perform well in this setting when used by MLHWs. None of the scabies diagnoses made by the experts were categorised as C2. This may be because such a pattern of scabies, atypical lesions or distribution and history of itch and contact with an affected individual, is rare in this setting. In this setting an individual diagnosed with suspected scabies due to an atypical clinical pattern is more likely to have another of the common pruritic skin disorders. The C2 category may prove useful in other settings where atypical clinical patterns have been reported[15].

Of the cases examined by MLHWs, the expert examiners identified burrows in 20 individuals (47.6% of cases). Burrows are challenging lesions to locate even for very experienced clinicians [15,16] and our study confirms, as reported elsewhere [11], that this is also the case for MLHWs. Four of the MLHWs failed to correctly locate any burrows, one MLHW detected burrows in three individuals. Whilst one MLHW did correctly identify burrows in eight individuals (40% of cases with burrows) they also noted burrows in eight participants where they had not been noted by the expert examiners, suggesting that this increased sensitivity came at the cost of a significant decrease in specificity. It is unlikely that detection of burrows will play a significant role in the diagnosis of scabies by MLHWs in communities with a high burden of disease. It is likely that the length of training for the MLHWs would needed to have been considerably longer to enable them to develop an improved ability to identify and locate burrows.

### Limitations

The MLHWs did not collect data concerning the distribution of lesions and we were not able to assess their performance in applying the B2 (male genitalia) criterion. We did not perform light microscopy of skin scrapings or a high-powered imaging device and so have not assessed two of the three criteria for confirmed scabies.

We were unable to assess the diagnostic accuracy of the MLHWs for impetigo and infected scabies due to the low number of cases seen. Performance of the non-expert examiners in the diagnostic accuracy study undertaken in the Solomon Islands showed they had a broadly similar performance with regards to the sensitivity and specificity of impetigo diagnosis compared with their diagnosis of scabies cases[11].

### Conclusions

The 2020 IACS Consensus Criteria for the Diagnosis of Scabies performed well when applied by dermatologists or MLHWs in this setting. The high level of diagnostic accuracy achieved by the MLHWs in our study indicates that this cadre of health professional is an effective resource for the diagnosis of scabies in LMICs. Further work to demonstrate the validity and reliability of the IACS criteria in other settings, rural and urban, community-based and in health care facilities and with other cadres of workers is needed. Scabies presents a significant health burden in many low resource settings and our study indicates that with short, focused training sessions, MLHWs could form a key component of scabies control strategies. To further inform control strategy development, it will be important to ensure MLHW training packages focus on the key diagnostic components of the IACS criteria and that attention is given to common differential diagnoses to reduce the likelihood of misdiagnosis. The design and delivery of sustainable educational interventions will need to be evaluated and appropriate individuals recruited to lead the training of MLHWs. Well trained MLHWs who are able to use the clinical diagnostic criteria will be key in the accurate assessment of the burden of scabies in LMICs and the implementation of strategies to control of scabies at the population level.

## Data Availability

Data are available in the supplementary materials

## Acknowledgements

The authors would like to thank the participants and members of the communities of Sinkor district Monrovia and Amos Ballah and Colette Baclene for their logistical support.

## Supporting information

Supplementary tables: Tables S1-S3

Supplementary data: Dataset S1 – Expert Examination, Dataset S2 – Non-Expert Examination

Supplementary checklist: Checklist S1 – STARRD checklist

